# Blood pressure trajectories through the first year postpartum following a hypertensive disorder of pregnancy

**DOI:** 10.1101/2023.10.13.23297033

**Authors:** Alisse Hauspurg, Samantha Bryan, Arun Jeyabalan, Esa M. Davis, Renee Hart, Jada Shirriel, Matthew Muldoon, Janet Catov

## Abstract

**Background:** Hypertensive disorders of pregnancy (HDP) are associated with future cardiovascular disease, predominantly through development of chronic hypertension, however the patterns of blood pressure recovery following a HDP are understudied. We sought to characterize the subtypes of hypertension (isolated systolic, isolated diastolic, systolic diastolic) and compare pregnancy and postpartum blood pressure trajectories among individuals with hypertensive disorders of pregnancy (HDP) who developed persistent hypertension at one year postpartum compared to individuals with normalization of blood pressure (BP).

**Methods:** We used data from a randomized controlled clinical trial of overweight and obese individuals with a physician adjudicated HDP conducted in the first year after delivery. Individuals with pre-pregnancy hypertension were excluded. Pregnancy BPs were obtained during prenatal visits, postpartum BPs were prospectively obtained through home BP monitoring, one week per month during the first year postpartum. Demographic characteristics and trajectories were compared based on whether or not individuals developed persistent hypertension (stage 1 or greater; systolic BP ≥130, diastolic BP ≥80 mmHg or use of anti-hypertensive medications) at one year. We further classified individuals with persistent hypertension as having isolated diastolic, isolated systolic and systolic diastolic hypertension. We used repeated BP measures to fit separate mixed-effects linear regression models for pregnancy and postpartum with participant identifier as random intercepts and weeks of pregnancy or months postpartum as a fixed effect expressed using restricted cubic splines. Models were compared using likelihood ratio test.

**Results:** We included 129 individuals who contributed a mean of 95.4 (95%CI 76.7-115.1) BP readings during and following pregnancy. In total, 75 individuals (58%) progressed to stage 1 or stage 2 hypertension by 1 year postpartum. At one-year postpartum, among those with persistent hypertension, 43 (69%) had isolated diastolic hypertension, 2 (3%) had isolated systolic hypertension and 17 (27%) had systolic-diastolic hypertension. Individuals with persistent hypertension were about 2 years older, delivered at earlier gestational ages and tended to have a higher BMI at one year postpartum compared to those with BP normalization. There were no differences in BP at first prenatal visit or BP trajectories during pregnancy. Individuals with persistent hypertension had a more adverse BP trajectory (p<0.01 for systolic and diastolic BP) in the first year postpartum. These differences persisted in multivariable models after adjustment for pre-pregnancy BMI and type of HDP (p<0.01 for systolic and diastolic BP).

**Conclusions:** Blood pressure trajectories in the first year postpartum, but not during pregnancy, may provide critical information for risk stratification after a HDP. In our study, a high proportion of individuals had ongoing hypertension, predominantly isolated diastolic hypertension. If confirmed in a larger cohort, this may provide insight into intervention development following a HDP.

CLINICAL TRIALS REGISTRATION URL: https://clinicaltrials.gov/ct2/show/NCT03749746

## INTRODUCTION

Hypertensive disorders of pregnancy (HDP), including gestational hypertension and preeclampsia complicate up to 10% of pregnancies in the United States with an increasing prevalence.^1^ The association of HDP with later-life cardiovascular disease (CVD) is well-established in diverse populations and is likely amplified in individuals with higher pre-pregnancy body mass index (BMI).^2–4^ This relationship is thought to be predominately driven by development of chronic hypertension. In the largest United States cohort followed from pregnancy to postpartum, the Heart Health Study, 37% of individuals with a hypertensive disorder of pregnancy developed chronic hypertension at 2-7 years after delivery.^5^

Despite the compelling evidence that the preponderance of HDP-associated CVD risk is linked to the development of chronic hypertension following pregnancy, the recovery of BP and transition to chronic hypertension remains understudied.^5,6^ Further, few studies have characterized the subtypes of hypertension (isolated systolic, isolated diastolic or both systolic and diastolic hypertension) associated with development of chronic hypertension following a hypertensive disorder. This may be particularly relevant to intervention development in this population as there are different physiologic drivers of hypertension which appear to vary by age with higher diastolic BP reflecting higher peripheral resistance in young adults whereas in older adults, higher diastolic BP reflects both arterial stiffness and peripheral resistance.^7,8^ As such, most contemporary intervention trials in the general population, including the SPRINT trial, target systolic blood pressure, which may be less relevant in a young population following a hypertensive disorder of pregnancy.^9^ Importantly, those with isolated diastolic hypertension have the lowest rate of awareness of their hypertension compared with people with isolated systolic hypertension.^10^

Our objectives were to (1) characterize clinical and obstetric risk factors and the subtypes of hypertension among those with persistent hypertension at one year postpartum and (2) compare pregnancy and postpartum blood pressure trajectories among individuals who developed persistent hypertension at one year postpartum compared to those with normalization of blood pressure.

## METHODS

### Description of parent study

We analyzed data collected through the Heart Health 4 New Moms (HH4NM) pilot randomized clinical trial. The details of the parent study have been previously published.^11^ Individuals with a singleton pregnancy complicated by a HDP (gestational HTN, preeclampsia without severe features, or preeclampsia with severe features) and no pre-pregnancy history of chronic hypertension were eligible for the parent trial. Briefly, Heart Health 4 New Moms is a prospective randomized trial (NCT03749746) of 148 individuals with a pre-pregnancy body mass index (BMI) in the overweight or obese range (body mass index [BMI] ≥25 kg/m^2^) with a hypertensive disorder of pregnancy who delivered between January 2019 and May 2021. Individuals were recruited in the postpartum period and randomized 1:1:1 to usual care, home BP monitoring program alone, or home BP monitoring plus an internet-based lifestyle program, HH4NM. All participants underwent two assessments: baseline (6 weeks to 6 months postpartum) and at study completion (8-12 months postpartum) after completing at least 6 months in the study. The diagnosis of a HDP was adjudicated by physicians (AH and AJ) using criteria from the American College of Obstetricians and Gynecologists (ACOG.

At the time of each research study visit, participants completed questionnaires and measured blood pressure at rest, following sitting for at least 5 minutes. Blood pressure measures were either collected by study staff (pre-pandemic) or self-collected with observation by study staff virtually using a secured Zoom room.. Blood pressures were measured three consecutive times using either an iHealth, Wireless Blood Pressure Monitor BP5 or an A&D UA-651 (A&D Medical; San Jose, California) automatic upper arm blood pressure monitor, both validated by Dabl Educational Trust and the British Society for Hypertension for use in pregnant and postpartum individuals. The mean of the three measures was used in analysis. The primary outcome of the trial was feasibility of a postpartum intervention study, and no differences were seen in weight or blood pressure between the three study arms, thus randomization groups were combined for this analysis.

### Description of analysis

We included individuals who completed both study visits. For our analysis, we classified individuals as having persistent postpartum hypertension if, at the time of the one-year postpartum research study visit, they had Stage 1 hypertension or greater (mean systolic BP ≥130mmHg or mean diastolic BP ≥80mmHg) or were taking antihypertensive medications. Stage 2 hypertension was defined as mean systolic BP ≥140mmHg or mean diastolic BP ≥90mmHg or treatment with antihypertensive medications. For the first objective, we compared clinical and obstetric outcomes among those with persistent hypertension. Among those with persistent hypertension, we then classified subtypes of hypertension as follows: isolated diastolic hypertension (IDH) was defined as mean diastolic BP ≥80mmHg with mean systolic BP <130 mmHg, isolated systolic hypertension (ISH) was defined as mean systolic BP ≥130mmHg with mean diastolic BP <80 mmHg. Combined systolic and diastolic hypertension (SDH) was defined as both systolic BP ≥130mmHg and diastolic BP ≥80mmHg. For the second objective, we used blood pressure collected at prenatal visits as part of routine clinical care for all participants with available data and data collected through postpartum home blood pressure monitoring among those randomized to the home blood pressure monitoring or home blood pressure monitoring + lifestyle intervention arms (n=81). For our home monitoring throughout the study period, participants were instructed by study staff on home measurement of blood pressure and were asked to measure blood pressure daily for one week out of each month, two measures in the morning and two measures in the evening, as previously described after study enrollment. In the immediate postpartum period, home BP data was collected through our institution’s clinical remote monitoring program (that included a mean of 19.4 ± 9.2 blood pressures during the first six weeks after delivery).^11,12^

### Statistical analysis

Statistical analysis was performed using Stata (StataCorp LLC, version 17.0, College Station, Texas, USA). Univariate analysis was conducted when indicated. Pearson’s chi-square tests were performed to analyze the differences between categorical variables. Two-sample two-sided t-tests and Mann-Whitney U tests were performed to analyze the differences between two groups of continuous, normally distributed and non-normally distributed variables, respectively. For all statistical tests, the alpha level was set at 0.05.

We then compared outpatient blood pressure trajectories during pregnancy and in the first year postpartum using separate mixed-effects regression models with weeks’ gestation or months postpartum as the timescale for analyses during pregnancy and postpartum, respectively. We used repeated blood pressure measures to fit mixed-effects linear regression models with each blood pressure measure as the outcome, participant identifier as random intercepts and time (weeks gestation or months postpartum) as a fixed effect expressed using restricted cubic splines with three knots in both models. The optimal number of knots were chosen comparing AIC and BIC between models. Mixed effects regression models were further adjusted for predefined covariates known to be associated with blood pressure including early pregnancy body mass index (BMI), maternal age and type of hypertensive disorder. Differences between the groups were tested via likelihood ratio test between models.

### Institutional approval

The protocol was approved by the Institutional Review Board of the University of Pittsburgh (STUDY18080007) and all participants provided written informed consent. Study data were collected and managed using REDCap electronic data capture tools hosted at the University of Pittsburgh. The study protocol was registered with Clinicaltrials.gov (NCT03749746) before any enrollment of participants.

## RESULTS

Overall, 129 of 148 participants enrolled in the parent trial completed the second study visit (87% retention; assessed at a mean of 10.9 ± 2.1 months after delivery). In our overall cohort, the majority of individuals had a diagnosis of preeclampsia (62%), with 30 (23%) requiring preterm delivery related to HDP diagnosis. In total, 75 individuals (58%) developed stage 1 or greater hypertension by one year postpartum and 28 (22%) developed stage 2 hypertension with 13 (10%) still requiring anti-hypertensive medications. Individuals with persistent hypertension (defined as stage 1 hypertension or greater) were similar to those with normalization of blood pressure in terms of age, race, pre-pregnancy body mass index (BMI), and family history (Table 1). As shown in Table 2, those with persistent hypertension delivered at earlier gestational ages (36.7±3.3 vs. 38.1 ±2.2 weeks; p=0.008) and were more likely to have a preterm delivery due to their hypertensive disorder (30% vs. 15%; p=0.05).

**Table 1.** Baseline demographics by hypertensive status at one year postpartum. Hypertensive was defined as systolic BP ≥130, diastolic BP ≥80 mmHg or use of anti-hypertensive medications.

| N(%), mean(SD) or median [IQR] | Normotensive | Hypertensive | p value |
| --- | --- | --- | --- |
|  | N=54 | N=75 |  |
| Age, years | 30.9 (5.3) | 32.6 (4.9) | 0.07 |
| Self-reported race |  |  |  |
| White | 46 (85) | 53 (71) | 0.10 |
| Black | 8 (15) | 19 (25) |  |
| Other (Asian, American Indian) | 0 (0) | 3 (4) |  |
| Pre-pregnancy / early pregnancy BMI, kg/m<sup>2</sup> | 30.5 [27.5, 34.3] | 31.2 [28.0, 35.6] | 0.35 |
| Health insurance type |  |  |  |
| Private / commercial insurance | 33 (61) | 53 (71) | 0.26 |
| Public insurance | 21 (39) | 22 (29) |  |
| Primiparous | 32 (62) | 35 (49) | 0.15 |
| Level of education |  |  |  |
| High school or less than high school | 12 (23) | 15 (21) |  |
| Some college or technical | 8 (26) | 23 (32) | 0.09 |
| 4 year college degree or higher | 33 (62) | 34 (47) |  |
| Family history of hypertension | 27 (50) | 42 (56) | 0.50 |
| Randomization arm |  |  |  |
| Control | 24 (44) | 24 (32) | 0.28 |
| Home BP monitoring alone | 14 (26) | 28 (37) |  |
| Home BP monitoring + HH4NM | 16 (41) | 23 (31) |  |

**Table 2.** Pregnancy characteristics by hypertensive status at one year postpartum. Hypertensive was defined as systolic BP ≥130, diastolic BP ≥80 mmHg or use of anti-hypertensive medications.

| N(%), mean(SD) or median [IQR] | Normotensive | Hypertensive | p value |
| --- | --- | --- | --- |
|  | N=54 | N=75 |  |
| Systolic BP at first prenatal visit, mmHg | 119 (10) | 120 (8) | 0.35 |
| Diastolic BP at first prenatal visit, mmHg | 74 (8) | 77 (6) | 0.10 |
| Gestational diabetes | 5 (9) | 10 (14) | 0.46 |
| Pregnancy diagnosis |  |  |  |
| Preeclampsia with severe features | 15 (28) | 31 (41) | 0.19 |
| Preeclampsia without severe features | 14 (26) | 20 (27) |  |
| Gestational hypertension | 25 (46) | 24 (32) |  |
| Gestational age at delivery | 38.1 (2.2) | 36.7 (3.3) | 0.008 |
| Preterm delivery due to HDP | 8 (15) | 22 (30) | 0.05 |
| Birthweight, grams | 3088 (641) | 2921 (840) | 0.22 |
| Small for gestational age | 5 (9) | 8 (11) | 0.86 |
| Gestational weight gain, pounds | 34.2 (19.4) | 30.1 (18.3) | 0.22 |
| Gestational weight gain above IOM recommendations<sup>a</sup> | 43 (80) | 52 (69) | 0.19 |
| Any breastfeeding (ever for this delivery) | 47 (87) | 58 (77) | 0.16 |

<sup>a</sup> Institute of Medicine (IOM) weight gain recommendations in pregnancy based on pre-pregnancy BMI (overweight: 15-25 pounds, obese: 11-20 pounds)

At the first study visit at approximately 10.4 ± 6.0 weeks postpartum, those who later progressed to persistent hypertension had a significantly higher systolic and diastolic blood pressure and were more likely to require anti-hypertensive medications (33% vs. 15%; p=0.02) compared to individuals with blood pressure normalization. Both groups had similar weight retention from pre-pregnancy and were equally likely to be lactating. (Table 3). At the final study visit at a mean of 10.9 ± 2.1 months, those with persistent hypertension had significantly higher systolic (127 ±10 vs. 114 ± 6; p<0.001) and diastolic (85 ±7 vs. 74 ± 4; p<0.001) blood pressure. Those with persistent hypertension had a higher BMI and more weight retention when compared to those with normalization of blood pressure.

**Table 3.** BP and weight at follow up visits by hypertensive status at one year postpartum. Hypertensive was defined as systolic BP ≥130, diastolic BP ≥80 mmHg or use of anti-hypertensive medications.

| N(%), mean(SD) or median [IQR] | Normotensive | Hypertensive | p value |
| --- | --- | --- | --- |
|  | N=54 | N=75 |  |
| Enrollment study visit |  |  |  |
| Time since delivery, months | 1.8 (1.1, 3.7) | 2.0 (1.4, 3.5) | 0.48 |
| Systolic BP, mmHg | 117.1 (10.8) | 128.4 (14.1) | <0.001 |
| Diastolic BP, mmHg | 77.4 (7.8) | 85.4 (9.9) | <0.001 |
| On anti-hypertensive medications | 8 (15) | 25 (33) | 0.02 |
| Stage 1 hypertension or greater | 26 (48) | 60 (80) | <0.001 |
| Weight change from pre-pregnancy, lbs | 9.2 (17.8) | 8.8 (19.5) | 0.90 |
| Body mass index, kg/m<sup>2</sup> | 31.7 [29.3, 35.9] | 32.7 [29.0, 37.6] | 0.55 |
| Breastfeeding currently | 34 (64) | 40 (56) | 0.38 |
| Follow up study visit |  |  |  |
| Time since delivery, months | 11.1 [9.2, 12.3] | 10.9 [8.8, 12.1] | 0.42 |
| Systolic BP, mmHg | 114 (6) | 127 (10) | <0.001 |
| Diastolic BP, mmHg | 74 (4) | 85 (7) | <0.001 |
| On anti-hypertensive medications | 0 | 13 (17) | 0.001 |
| Weight change from pre-pregnancy, lbs. | 7.3 (20.7) | 11.3 (20.5) | 0.29 |
| Body mass index, kg/m<sup>2</sup> | 31.8 [27.9, 36.0] | 33.1 [29.1, 40.1] | 0.10 |
| Breastfeeding currently | 19 (35) | 22 (29) | 0.48 |

Next, we sought to characterize subtypes of hypertension among those with persistent hypertension. At one-year postpartum, among those with persistent hypertension not on anti-hypertensive therapy (n=62), 43 (69%) had isolated diastolic hypertension, 2 (3%) had isolated systolic hypertension and 17 (27%) had systolic and diastolic hypertension. Among those on anti-hypertensive medications (n=13), 5 (38%) were normotensive (<130/80 mmHg), 5 (38%) had isolated diastolic hypertension, none had isolated systolic hypertension and 3 (23%) had systolic and diastolic hypertension (Figure 1).

**Figure 1.**
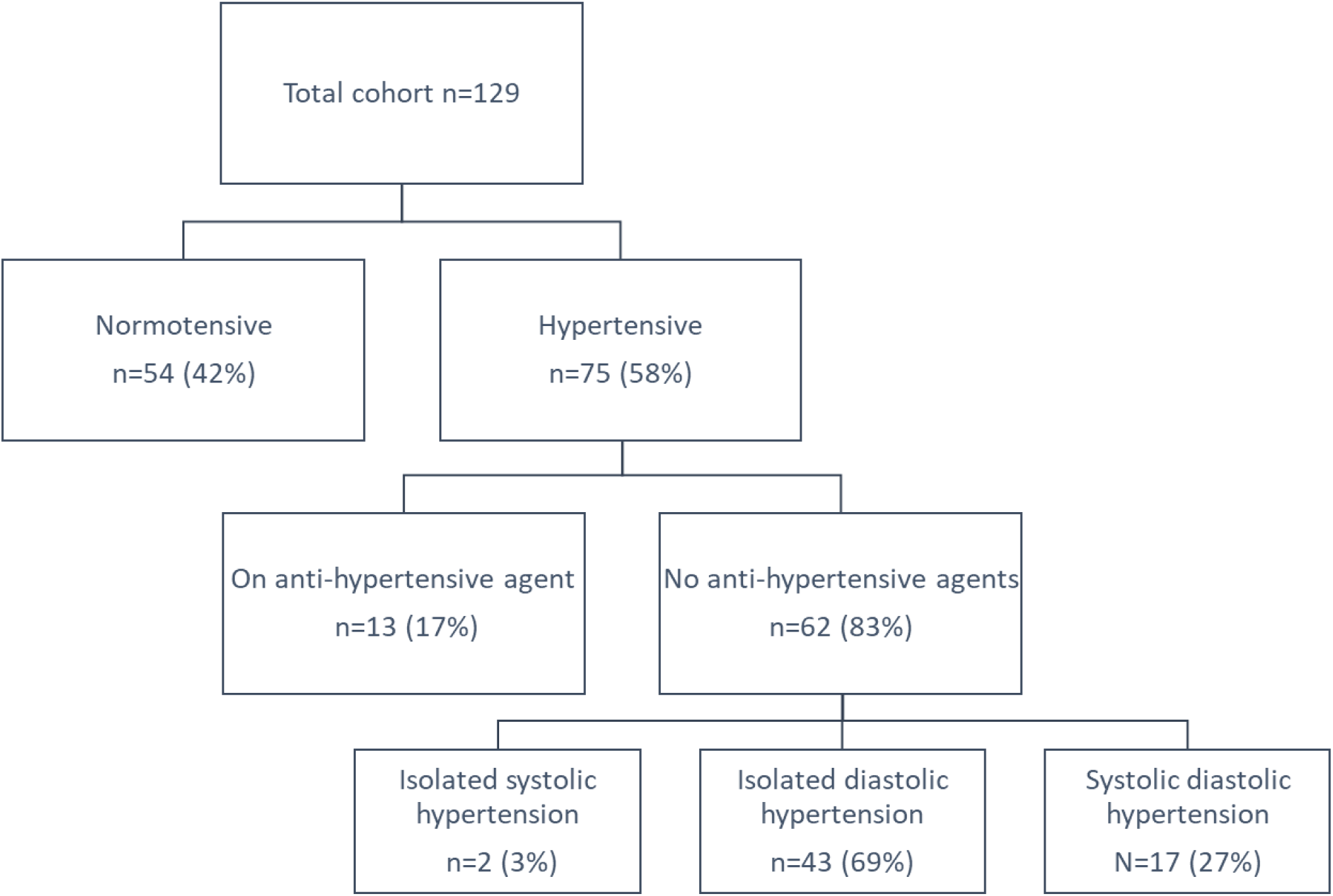
Subtypes of hypertension at one-year postpartum. Isolated diastolic hypertension was defined as mean diastolic BP ≥80mmHg with mean systolic BP <130 mmHg, isolated systolic hypertension was defined as mean systolic BP ≥130mmHg with mean diastolic BP <80 mmHg. Combined systolic and diastolic hypertension was defined as both systolic BP ≥130mmHg and diastolic BP ≥80mmHg.

Finally, we sought to evaluate differences in blood pressure trajectories during pregnancy and postpartum between those with persistent hypertension compared to those with normal BP. Across both groups, 993 blood pressures measured at prenatal visits in 108 individuals were available for analysis. There were no differences in systolic (120 ±8 vs. 119±10; p=0.35) or diastolic (77±6 vs. 74±8; p=0.10) blood pressure at the initial prenatal visit between groups. Similarly, systolic (p=0.20) and diastolic (p=0.20) blood pressure trajectories during pregnancy did not differ by group (Figure 2a, 2b). Among individuals in the home blood pressure monitoring groups (n=81; 30 [37%] with BP normalization and 51 [63%] with persistent hypertension), we then examined differences in postpartum blood pressure trajectories. In total, 14,177 postpartum home blood pressures were included. Individuals with persistent hypertension had a more adverse systolic (p<0.01) and diastolic (p<0.01) blood pressure trajectory in the first year postpartum. These differences persisted in multivariable models after adjustment for age, pre-pregnancy BMI and type of HDP (p<0.01 for systolic and diastolic BP).

**Figure 2.**
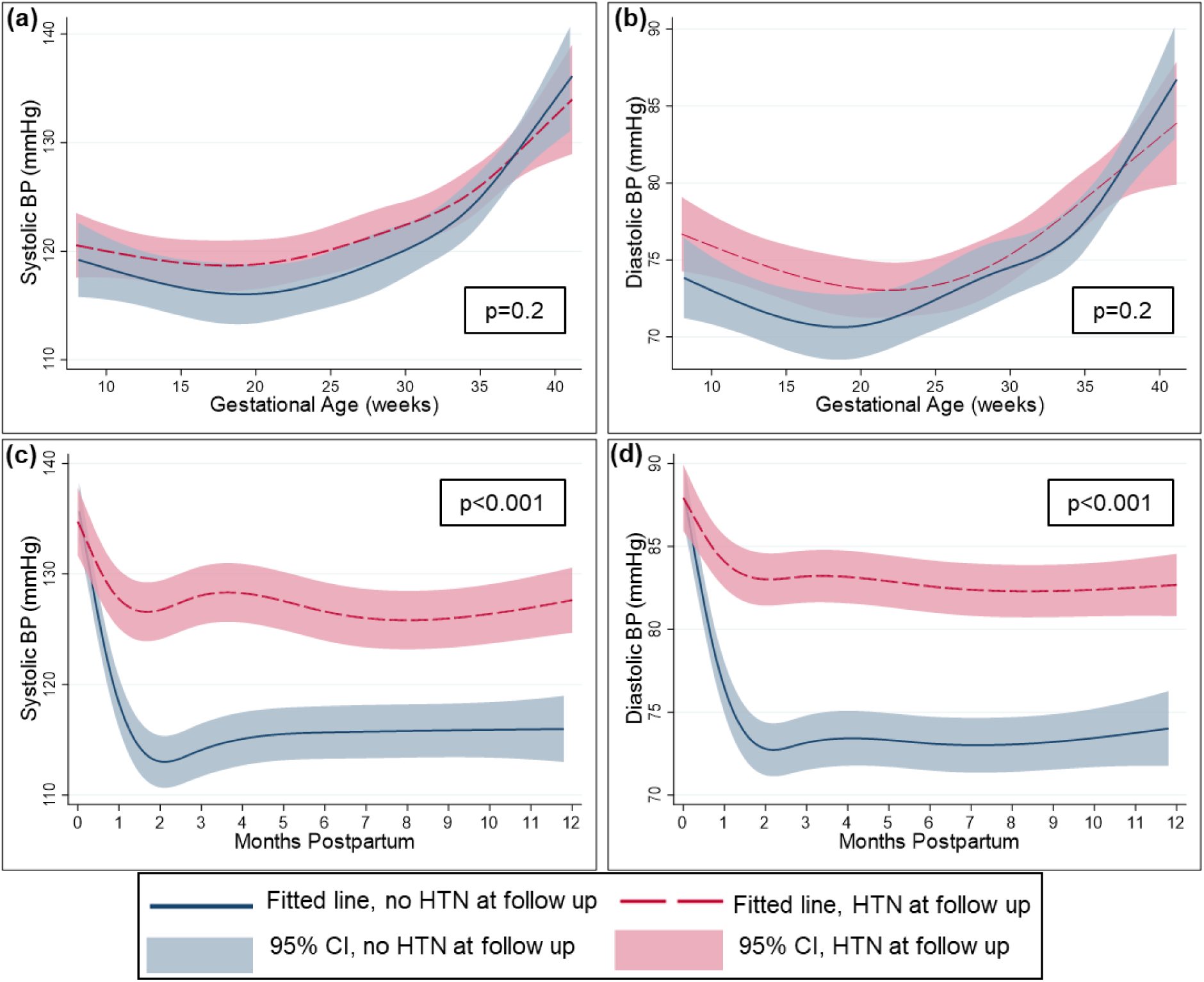
Systolic and diastolic blood pressure trajectories during pregnancy (panels a, b) and postpartum (panels c, d) by hypertensive status at one year postpartum. Hypertensive was defined as systolic BP ≥130, diastolic BP ≥80 mmHg or use of anti-hypertensive medications.

## DISCUSSION

### Main findings

In this analysis, using prospectively collected data with high participant retention, we longitudinally describe blood pressure trajectories and subtypes through one-year postpartum following a hypertensive disorder of pregnancy. Despite no differences in prenatal blood pressure patterns, blood pressure trajectories in the first year postpartum differed significantly among those with persistent hypertension compared to those with BP normalization, suggesting that the BP trajectory may provide critical information for risk stratification after a HDP. In our study, a high proportion of individuals had ongoing hypertension, suggesting the importance of home BP monitoring for additional postpartum risk stratification following a HDP. Finally, among those with persistent hypertension, the majority of hypertension was isolated diastolic, which may be particularly important for intervention development, targeted treatment and future clinical care guidelines.

### Interpretation

The clinical approach in obstetrics has been that the pathogenesis of preeclampsia is resolved with delivery. This hands-off approach to management after delivery has likely contributed to the increasing maternal morbidity and mortality seen in the postpartum period and has also led to a poor understanding of blood pressure recovery following a hypertensive disorder of pregnancy.^13^ Recent evidence suggests the immediate postpartum period is a critical time for cardiovascular remodeling, with implications for long-term maternal health.^14,15^ Small RCTs have demonstrated that improved management of hypertension immediately postpartum leads to longer-term BP benefits.^14–16^ In the SNAP-HT trial, done in the United Kingdom, self-managed postpartum hypertension (among individuals with gestational hypertension or preeclampsia) with home monitoring and tight BP control led to improved diastolic BP at 6 months postpartum that was sustained through 3-4 years after delivery in n=62 women. A similar proportion of individuals were on anti-hypertensive medications at 6 months postpartum across both arms, but tighter BP control during the critical immediate postpartum period led to lower BP long-term (-4.5 mmHg office diastolic BP at 6 months and -6.8 mmHg ambulatory diastolic BP at 3-4 years). Another small study randomized individuals with preterm preeclampsia to postpartum enalapril and demonstrated improved left ventricular remodeling at 6 months postpartum when compared to individuals randomized to placebo.^14^ These studies suggest that the immediate postpartum period may be particularly important for cardiovascular remodeling and that interventions in this period may improve both short and long-term cardiovascular risk. Our findings support the importance of blood pressure patterns in the first few weeks after delivery as more informative than pregnancy BPs for longer-term risk of hypertension, emphasizing the potential importance of ongoing home BP monitoring following a HDP. Home monitoring of BP both during pregnancy and postpartum has been previously demonstrated to be feasible and acceptable to pregnant and postpartum individuals and improves ascertainment of blood pressure in the postpartum period.^12,17,18^

One major limitation to interventions beyond the immediate postpartum period is access to care. For individuals with public health insurance, which comprises over 40% of deliveries, in many states within the United States, Medicaid coverage lasts only through sixty days postpartum.^19^ The high rates of ongoing hypertension in our population highlight the critical need for universal Medicaid expansion to at least one year postpartum. Pregnancy can be viewed as a window to future cardiovascular health, and as such, the first year postpartum, likely provides additional insights into long-term health and pre-conception health for a potential subsequent pregnancy. Individuals identifying as female remain underrepresented in cardiovascular clinical trials relative to their percentage of the population and disease burden.^20–22^ The urgency of these trends is amplified by the fact that chronic hypertension is a condition that increases more rapidly during the reproductive years with worse morbidity in females compared with males.^23^ Compared to males, females are diagnosed later, are less likely to receive guideline-concordant care and overall quality of care is significantly lower.^24,25^ Collectively, these findings translate to stagnant mortality rates from CVD in individuals identifying as women, particularly young women, and emphasize the importance of approaches that are centered on primary prevention.^24,26^ Our data supports the importance of efforts to improve long-term CVD by focusing on the first year after delivery as a critical opportunity for primary prevention to improve short and long-term health.

Our finding that the majority of hypertension at one year postpartum in our population is isolated diastolic hypertension further underscores the importance of provider and patient education about CV risk following a hypertensive pregnancy. Those with isolated diastolic hypertension (IDH) have the lowest rates of awareness of their hypertension and are less likely to receive a hypertension diagnosis compared to those with elevated systolic blood pressures. In studies that are limited to young adults, IDH has been shown to be associated with adverse outcomes, including CVD events, heart failure, atrial fibrillation and chronic kidney disease.^27–29^ Despite this, most BP guidelines do not specifically address IDH as a distinct phenotype and provide no guidance for its management. Data from a larger, diverse cohort of describing hypertensive subtypes in the years after pregnancy may help inform the design and development of future intervention trials and ultimately clinical guidelines in the postpartum period.

### Strengths and Limitations

Our study is strengthened by the prospective nature with high adherence to study protocols and participant retention of 87% up to one-year postpartum. We collected a large number of blood pressures during pregnancy and in the postpartum period allowing longitudinal exploration of blood pressure patterns. Our definition of persistent hypertension was made from measurement of blood pressure in triplicate; however, this was from a single study visit. Future studies would be strengthened by diagnosing hypertension based on BP measured at more than one research study visit. We are further limited by the small sample size in a single geographic area, limiting its generalizability. Eligibility for the parent study required individuals to have a device with internet access, which may have limited the economic diversity of individuals eligible to participate. Finally, we included only overweight and obese individuals, and while close to 70% of reproductive-aged women now fall into these categories, our findings may not be applicable to underweight or normal weight individuals.^30^

Overall, in this study, we demonstrate that individuals with persistent hypertension at one year postpartum had similar BP trajectories during pregnancy but a blunted BP recovery postpartum compared to individuals with normalization of BP. We demonstrate a high prevalence of persistent hypertension, predominantly isolated diastolic hypertension. Our findings emphasize that the blood pressure trajectory in the first year postpartum may provide critical information for risk stratification after a HDP and the importance of interventions targeting diastolic hypertension.

## Data Availability

Data that support the findings of this study are available from the corresponding author upon reasonable request and following IRB rules and privacy regulations.

